# ‘Navigating a lonely road to adulthood with an ever-changing map’; A Framework and Case Study Protocol Exploring the Transition to Adulthood, Mental Health Literacy and Occupational Balance among Swedish Young Adults

**DOI:** 10.1101/2025.06.19.25329972

**Authors:** Martin Karaba Bäckström, Sonya Girdler, Ulf Jonsson, Carita Håkansson, Annika Lexén

**Author notes:** Corresponding author: <u>Martin Karaba Bäckström,</u> ‘Mental Health, Activity and Participation’ Department of Health Sciences, Lund University Sölvegatan 19, 223 62, Lund, Sweden. Competing interests None declared. Funding This research is funded by FORTE (grant number: 2023-00254), the Swedish Research Council for Health, Working life and Welfare. Availability of data and materials Original research data was neither collected or generated for this publication.

## Abstract

**Background:** Recent reports point to a global and Swedish national mental health among young adults. It is widely recognised that this phase of ‘emerging adulthood’ involves many challenges, stressors and uncertainties that can negatively impact mental health and well-being. In addition, these strains may be exacerbated by rapid contemporary changes in technological, societal and sociocultural contexts. Developing and tailoring interventions bolstering the mental well-being of young adults requires deeper insights into their lived experiences, self-perceived strengths and challenges.

**Aim:** This paper has two interconnected purposes: Firstly, presenting relevant theories and theoretical perspectives at the individual, group and societal levels providing a rationale for theorised study propositions underpinning a situational analysis of Swedish young adults. Secondly, building on this theoretical foundation, the aim is to describe a protocol for a case study and situational analysis examining how Swedish young adults from Generation Z experience their transition to adulthood, its impacts on their mental health, describing their mental health literacy, occupational balance, and support needs.

**Method:** This case study protocol outlines a holistic, single-case study design employing a flexible approach. Data will be collected in Sweden through focus groups and individual interviews with stakeholders, young adults, and their parents. Existing and available mental health and well-being promotion in Sweden will be mapped through online searches. The various data sources will initially be analysed separately using thematic analysis. Subsequently the subthemes and themes will be jointly analysed across participant groups using pattern matching in workshop formats.

**Conclusions:** The results of this case-study are expected to generate new knowledge of the reasons underpinning the increase in mental ill-health among young adults in Sweden. This knowledge will subsequently inform the future co-production and core design features of a digital mental e-health intervention aimed at promoting mental well-being among Swedish young adults aged 18–29.

## Introduction

Internationally and in Sweden, there is evidence of a continuing mental health crisis among young adults [1,2]. The prevalence of both short and long-term mental ill-health is rising, with suicide remaining the third leading global cause of death in this age group, and the second leading cause of death in Sweden [1,3,4]. Anxiety, stress, worry and sleep disturbances are common experiences in the daily lives of Swedish young adults [2]. Further, the onset of mental ill-health during young adulthood accounts for a substantial proportion of disability-adjusted life-years, years lived with disability, and years of life lost [5–7]. Today’s generation of young adults (aged 18-29 years), commonly referred to as ‘Generation Z’ [8], face not only the development challenges inherent in ‘emerging adulthood’, the life phase between 18-29 characterised by uncertainties, possibilities and finding one’s role in society [9,10], but also evolving societal stressors and megatrends such as a rapidly evolving job market and the negative consequences of connectivity and digital technology [11]. While Swedish young adults increasingly report escalating mental ill-health and challenges in navigating this critical life phase [2], the link between mental health and well-being during emerging adulthood and broader societal and sociocultural factors remains largely underexplored [10].

The high rates of mental ill-health among young adults represents one of the most pressing threats to individual, community, and societal well-being, development, and economic stability on a global scale [1,3]. While research highlights that investments in mental health and well-being promotion, prevention, and intervention for young adults are not only necessary, but also cost-effective [1,3,7], the efficacy and positive outcomes of these initiatives depend on a thorough understanding of the unique living conditions, needs, and challenges faced by young adults today [11,12]. This is particularly important given Generation Z is commonly described as a generation whose lifestyles differ significantly from those of previous generations [13], highlighting the importance of exploring their generational traits, perspectives on mental health, and experiences of emerging adulthood. Currently, no situational analysis is available examining the perspectives of Swedish Generation Z on mental health and well-being during this transitional phase.

### Framework and case study protocol

This paper has two interconnected purposes: Firstly, presenting relevant theories and theoretical perspectives at the individual, group and societal levels providing a rationale for theorised study propositions underpinning a situational analysis of Swedish young adults. Secondly, building on this theoretical foundation, the aim is to describe a protocol for a case study and situational analysis examining how Swedish young adults from Generation Z experience their transition to adulthood, its impacts on their mental health, describing their mental health literacy, occupational balance, and support needs. The framework and the associated study propositions (see Table 1) were member-checked and evaluated during a two-hour digital workshop with an expert panel of five Swedish young adults aged 18–29. Held in April 2025, the workshop aimed to gather feedback on the proposed theories, seeking further suggestions, obtaining feedback on the framework’s name and the relevance of the study propositions to the participants’ lived experiences. Additionally, two young adults who were members of the expert reference panel for the study, provided written feedback contributing to the development of the theoretical framework.

**Table 1.**
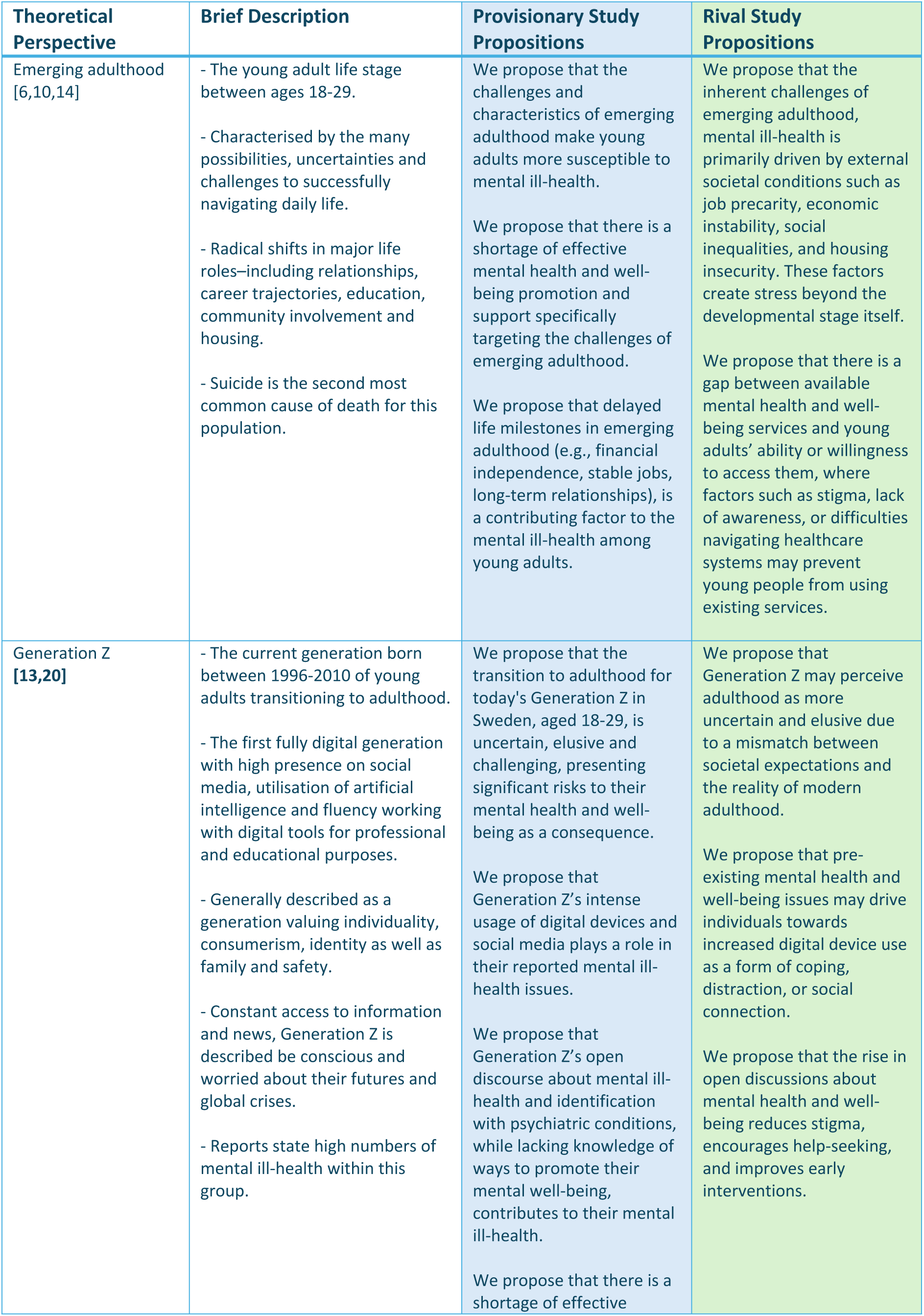
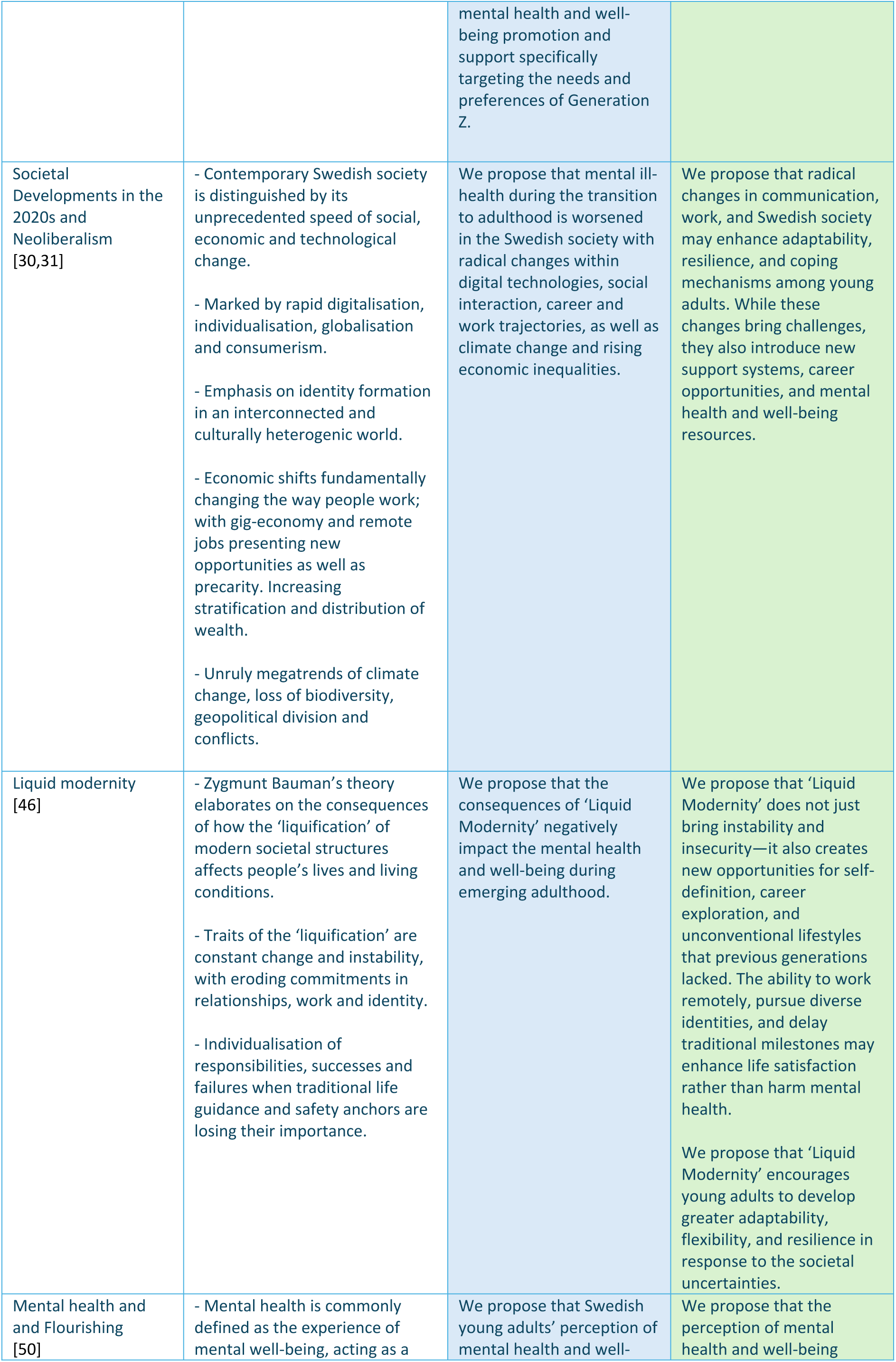
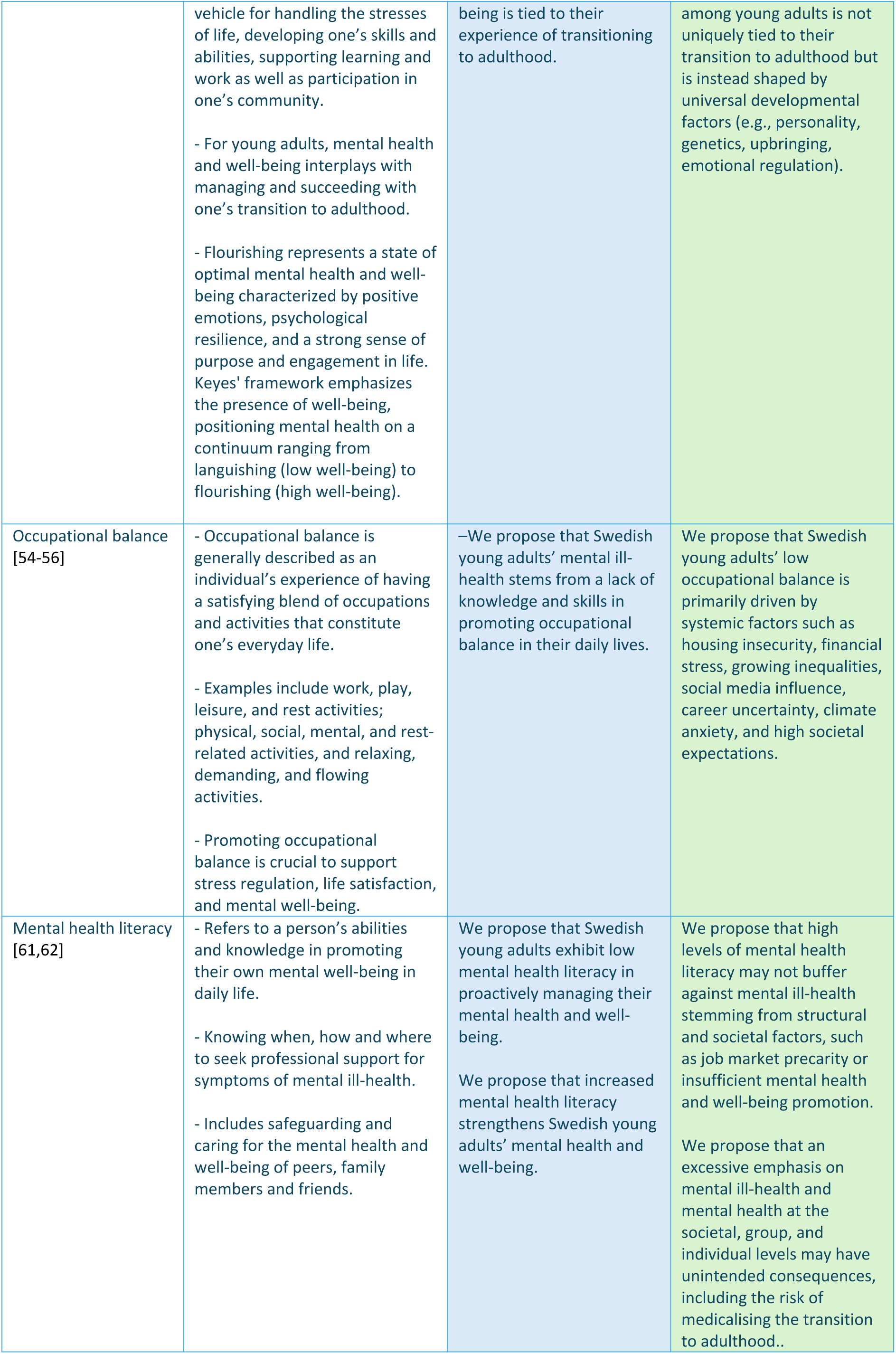
Theoretical perspectives and theoretical study propositions of a framework exploring emerging adulthood in Sweden today and its implications for mental health.

#### ‘Navigating a lonely road to adulthood with an ever-changing map’: A framework for understanding today’s challenges to the mental well-being of Swedish young adults

This framework seeks to integrate perspectives and theories at the individual, group, and societal levels contextualising the ongoing crisis of mental ill-health among Swedish young adults in Generation Z. ‘Navigating the journey to adulthood with an ever-changing map’ symbolises the complex interplay of factors that contribute to the mental health and well-being of this generation. The framework incorporates several theoretical perspectives, grouped as follows: ‘Emerging adulthood’ and ‘Generation Z’– contextualising the young adult life phase and the defining characteristics of today’s young adults. Societal developments in the 2020s – providing a contextual backdrop through the theories of ‘Neoliberalism’ and ‘Liquid Modernity’. Finally, theories of mental health, ill-health and well-being, occupational balance and mental health literacy providing a foundation for understanding the factors that promote or hinder mental well-being during emerging adulthood.

### Emerging Adulthood and Generation Z

‘Emerging adulthood’ (18-29 years) is a pivotal and distinct life stage during which individuals transition from adolescence to independent adulthood [14]. It is characterised by significant biological, psychological, and neurodevelopmental changes, including ongoing brain maturation and emotional regulation [14]. This period is marked by significant transformations across multiple life domains, including education, employment, independent living, relationships, and community engagement. These transitions often present both opportunities and stressors that can contribute both to mental well-being and deteriorating mental ill-health [6,10]. A defining feature of emerging adulthood is the experience of being “in-between”, requiring constant adaptation to new roles, activities and relationships while simultaneously leaving behind established familiar safety nets [14].

The wide range of choices available and demands of the decision-making needed to accomplish and ‘fill the shoes’ of various adult roles have been linked to increased levels of anxiety and stress among young adults [6,15]. Simultaneously, the 2020’s are characterised by ‘neoliberalist values’ of individual choice and success, constant digital connectivity and social media usage, believed to exacerbate young adults’ fear of making the ‘wrong’ choices regarding their futures [16,17]. While previous studies on emerging adulthood in Sweden have largely focused on identity development [18,19], little is known about how young adults today perceive the transition adulthood and its implications for their mental health. Given the complexity and demands of this life stage, young adults are at heightened risk of experiencing elevated stress levels, anxiety, and mental ill-health [6,10].

‘Generation Z’ [13,20] (born between 1996 and 2010) is the group of young adults currently transitioning to independent adulthood. This generation is characterised by its deep integration with digital technology, individualistic ideals, and emphasis on identity expression [13,20]. Compared to previous generations, Generation Z exhibits greater differences from its immediate predecessors than any other generational shift [20]. Social media and smartphone use play a central role in their daily lives, serving as a source of both stress as well as relaxation, community connection, contributing to identity formation [13,21,22]. Although young adults today are more connected than ever through digital technology and intense use of social media, these connections are often experienced as superficial, alienating, and can lead to unfavourable comparisons, reduced self-esteem, loneliness, and self-worth [22,23]. One notable example of an online community that superficially appears supportive, but actually poses risks to the mental well-being of young adults, is the internet phenomenon known as looksmaxxing [24]. Looksmaxxing refers to efforts to maximise one’s physical appearance and lifestyle in order to become more conventionally attractive, with the aim of gaining social benefits and increasing the likelihood of intimate relationships with men or women [24]. This practice is often associated with global online ‘manosphere’ and ‘incel’ communities, which are often misogynistic and influenced by male supremacy ideologies [24,25]. The number of young men identifying with these movements is increasing, with Sweden being noted as having a high estimated prevalence of members per capita [25,26]. A recent study [24] analysing 8,072 comments on looksmaxxing forums found that self-improvement efforts can involve dangerous at-home surgical procedures, such as ‘bonesmashing’ to alter the jawline. The study also highlight the use of a demoralising, hypermasculine jargon that undermines self-worth and self-esteem, and can encourage suicidal ideation—commonly expressed with phrases like ‘to rope’ or ‘it’s over’ when a man is perceived as beyond help [24].

Generation Z has been described as valuing consumerism, materialism and economic wealth as primary goals for a successful adult life, values associated with concerns about the future and a generally pessimistic outlook on societal developments [20]. In terms of mental health, studies report an increasing prevalence of psychiatric diagnoses and mental ill-health in this age group [1,2,13]. Research presents a contradictory picture of whether young adults are more open in sharing their experiences of mental ill-health. While tendencies toward greater openness have been observed [8,13], 60% of Swedish young adults aged 18-29 report difficulties in doing so [27]. Recent findings [8,28] also indicate that many young adults turn to social media for mental health and well-being guidance and support, a practice that entails both the risk of misinformation and the potential for peer support and connection.

### Societal Developments in the 2020s, Neoliberalism and Liquid Modernity

Globally, the 2020s are characterised by rapid structural changes and megatrends that impact young adults’ mental health and well-being [29], including accelerated digitalisation, increasing individualisation, socioeconomic inequalities, and geopolitical instability [30,31]. Illustrations of these shifts are climate change and loss of biodiversity, technological developments in the fields of generative artificial intelligence, information and communication technologies, as well as increasing economic inequalities and stratifications, geopolitical conflicts and wars. The COVID-19 pandemic further exacerbated socioeconomic disparities, highlighting deep-seated social vulnerabilities and worsening mental health and well-being among young adults [30,31]. One notable consequence of these global challenges is climate anxiety, a growing psychological response to the existential threat of climate change. This phenomenon is increasingly affecting young adults, leading to heightened stress, feelings of helplessness, and uncertainty about the future [32]. Research suggests that many young adults experience eco-anxiety as a persistent worry about environmental degradation and its long-term consequences, exacerbating existing mental health struggles, contributing to a sense of a lack of control over their future [33].

While in contemporary Sweden most young adults report good mental health [2,34] many still face considerable challenges to their mental health and well-being during their transition to adulthood, particularly those from minority, marginalised or disadvantaged groups. For instance, young adult Samí (Sweden’s indigenous population) report worse mental health and well-being than their majority counterparts, with higher incidences of depression and suicidal thoughts or attempts [35]. These trends are also seen among Swedish LGBTQ+ young adults and persons with disabilities, who also experience lower rates of overall mental well-being and health [36,37]. Among all Swedes, social isolation is another pressing issue, with nearly 40% of individuals aged 16–29 reporting ill-health stemming from involuntary loneliness, and over 25% of young men lacking emotional support in their daily lives [38]. Further, 74.7 % of young women and 44.7 % of young men report experiencing anxiety, worry, and distress in daily life [34].

Several thresholds are currently hindering Swedish young adults’ transition to adult roles, including socioeconomic gradients relating to quality of housing, education and social support systems impacting their mental health and transition to adulthood [37]. Factors such as their parents’ educational levels and income, immigrant background, where a young adult has grown up and gone to school, play significant roles in their future opportunities and resources for mental ill-health or well-being [37]. The majority of Swedish young adults reside in municipalities where affordable housing is scarce [39], while the Swedish youth unemployment rates remain at a national high of 24.3% [40]. Financial instability is a growing concern with increasing levels of debt, particularly related to often necessary student loans and consumer credit [41,42]. Many young adults struggle with financial literacy and access to stable income, leading to potential long-term economic vulnerability [42].

Additionally, young adults who fail to complete upper secondary education experience significant disadvantage in the labour market, reporting higher unemployment rates, lower income levels, and reduced job security [43]. For students in higher education, stress and mental ill-health concerns are prevalent. Academic pressure, financial strain, and uncertainty about future employment are all contributing to heightened distress [27]. Reports indicate that a substantial proportion of university students experience symptoms of burnout, sleep disturbances, and anxiety due to the demands of their studies and economic pressures [41].

Similar symptoms are also apparent among young adults commencing their careers, and are the primary drivers of short and long-term sick leave in this group [44]. Young women between 25-29 are especially vulnerable due to the combined gendered roles of working professionally and unpaid work at home [44]. Working life is often experienced as stressful and finding time for rest during and outside working hours is as a major challenge across several industries and professions [44].

‘Neoliberalism’ is an economic and political ideology that emphasizes freedom of choice, individual responsibility, privatisation, free markets and the minimisation of state intervention in society and welfare [16]. This ideology shifts the burden of success or failure onto the individual, fostering an environment of intense competition and self-optimization. For Generation Z, currently transitioning to adulthood and in the midst of establishing their careers, neoliberalism ideals increase their risk of loneliness, feelings of financial instability, and job precarity, contributing to pressure to enhance their personal and professional profiles in an increasingly competitive landscape [16,17]. The erosion of stable career paths, rising student debt, and unaffordable housing create stress and anxiety [17]. Further, the strong emphasis on personal responsibility can lead to feelings of failure and low self-esteem when young adults struggle to meet societal expectations [45]. The emerging cultures of digital hustle, gig work, and social media-driven self-branding can be seen as direct consequences of a neoliberal economy, potentially negatively impacting the mental well-being of young adults [16].

Bauman’s theory of ‘Liquid Modernity’ [46] describes a contemporary world in which traditional markers of stability, such as long-term employment, lifelong relationships, and predictable life paths, are increasingly elusive. Bauman argued that liquid modernity shifts responsibility for success and failure onto the individual, detaching them from collective systems of support. Bauman’s theory [46] can be particularly relevant for understanding the mental health and well-being challenges faced by Generation Z. For example, the uncertainty surrounding life trajectories can generate immense pressure to constantly prove oneself, thereby increasing the risk of stress and anxiety [1,18].

### Mental Health, Ill-health and Flourishing

The World Health Organisation defines [3] ‘mental health’ as a state of mental well-being, enabling individuals to cope with life’s stressors, fulfil their potential, succeed in learning and work, and contribute positively to their community. Conversely, ‘mental ill-health’ encompasses a wide range of short and long-term issues, psychosocial disabilities and other mental states that cause significant distress, impair functioning, or pose a risk of self-harm [3]

Several notable scholars and researchers have contributed to further conceptualizations of mental health and well-being [47–49]. For this framework, we have chosen Keyes theory of ‘flourishing’ as a guiding metaphor on how mental well-being can be understood from the perspective of transitioning to adulthood [50]. Flourishing, as defined by Keyes, represents a state of optimal mental health and well-being characterized by positive emotions, psychological resilience, and a strong sense of purpose and engagement in life [51,52].

Unlike traditional models that primarily focus on the absence of mental illness, Keyes’ framework emphasizes the presence of well-being, positioning mental health on a continuum ranging from languishing (low well-being) to flourishing (high well-being) and can vary on a day to day basis [50]. One study based on interviews with young adults about the concept of flourishing found that they associated this term with words such as ‘thriving’, ‘fullest potential’ and blooming flowers [53] and considered it a relevant concept when discussing mental well-being.

This theory is particularly relevant during young adulthood, given this life stage involves significant developmental, social, and occupational transitions that can influence mental well-being [6]. Factors such as identity formation, educational and career uncertainty, social belonging, and autonomy-seeking behaviours play a crucial role in determining whether an individual thrives or struggles during this period. By integrating the concept of flourishing into our framework, we aim to explore how young adults experience well-being amidst the challenges and opportunities that define emerging adulthood.

### Occupational Balance and Mental Health Literacy

‘Occupational balance’ refers to an individual’s ability to maintain a subjectively satisfying variance and dosage of meaningful activities in one’s daily life [54–56]. This includes the distribution of time and energy spent on obligatory and discretionary activities. Occupational balance can also be understood as a variety activities promoting feelings of relaxation, accomplishment and flow [54–56], which contribute to an individual’s mental well-being, ability to handle stress and resilience against mental ill-health [55–57]. Generation Z has been described as showing signs of low occupational balance, reporting difficulties in maintaining a sustainable balance between work and study-related activities and those that promote recovery and enhanced quality of life. This imbalance poses a significant risk in increasing their feelings stress and experiencing mental ill-health [22,58]. While further research is needed to understand the association between technology use and mental ill-health among generation Z it has been noted that excessive smartphone and social media use may also be factors contributing to decreased occupational balance among young adults [59,60].

‘Mental health literacy’ [61,62] refers to an individual’s ability to promote their mental well-being while recognising the early signs of deteriorating mental health. It includes understanding both the protective and detrimental factors influencing mental health, knowledge of mental health conditions, and the ability to provide support to others experiencing mental ill-health [61,62]. Equally important is developing self-care skills that help individuals manage stress and emotional challenges. Improving mental health literacy at the individual, group, and community levels is essential for reducing stigma, fostering empathy, and promoting mental well-being [61,62]. Additionally, higher levels of mental health literacy facilitate early intervention by equipping individuals with the knowledge of when, how and where to seek professional or community-based support [63].

While limited, available research examining the mental health literacy of Generation Z suggests they have relatively low levels of awareness and preparedness for managing their mental health and well-being. Their scant understanding of practical ways to promote their mental well-being in their everyday lives is particularly concerning [2,64]. Reflecting the rise of social media in recent decades, Generation Z demonstrates a greater propensity than previous generations to disclose and engage in discourse surrounding mental health and well-being difficulties [13,20,65]. Notably, there is a lack of research examining the mental health literacy of Swedish young adults.

### Theoretical study propositions originating from the framework

Contemporary Swedish society encompasses a range of structural and social processes that influence the mental health and well-being of young adults. Sustained engagement with digital platforms contributes to burnout and reduces in-person socialization, while exerting a pressure to maintain a curated online persona contributing to anxiety and self-esteem issues [22,66,67]. Economic stressors, such as competitive education systems and precarious job markets, create widespread experiences of stress, anxiety, and feelings of inadequacy. The erosion of traditional social safety nets (such as stable career paths and strong community support systems) has left young adults navigating their transition to adulthood in the context of liquid modernity, where uncertainty is a defining characteristic [13,20,46]. The rates of mental ill-health and suicide among young adults underscore the urgent need for innovative and evidence-based efforts to promote mental well-being. Understanding how young adults experience their transition to adulthood and how they manage their mental health and well-being is critical for developing effective promotion interventions.

### Case study protocol

Secondly, building on this theoretical foundation, the aim is to describe a protocol for a case study and situational analysis examining how Swedish young adults from Generation Z experience their transition to adulthood, its impacts on their mental health, describing their mental health literacy, occupational balance, and support needs. The following sections describe the planned execution of the case study, using Yin’s [68] recommended headings for case study protocols. This protocol also adheres to the ‘Standards for Reporting Qualitative Research’ [69] (SRQR) guidelines, ensuring the scientific rigour of the forthcoming case study.

#### Overview of the case study

The main goal of this situational analysis is to provide insights for producing a digital application that promotes mental health literacy, peer support and well-being in collaboration with Swedish young adults and other stakeholders [22]. Although multiple digital mental health and well-being interventions have shown efficacy in treating various mental ill-health symptoms, sustaining user engagement remains a challenge [70]. One strategy to address this issue is to utilise co-production models where the end users’ lived experiences are placed at the core of the design processes [71]. Examples of such models are the Medical Research Council’s ‘Framework for Developing and Evaluating Complex Interventions’ [72], principles of ‘Design Thinking’ [73] and ‘Participatory Design of evidence-based online youth mental health promotion, intervention and treatment’[71]. These models provide a structured approach to creating solutions for prevention, health promotion, and care—from idea generation to implementation.

This case study focuses on the initial design stage, commonly referred to as ‘needs assessment’ phase or, ‘identify’ and ‘define’ [71–73]. This stage aims to understand the target group’s experiences, circumstances, and unmet needs related to mental health and well-beingand well-being. By gaining a deep understanding of the challenges and strengths experienced by Swedish young adults regarding their mental health, the subsequent digital mental health and well-being intervention is more likely to be aligned with their needs and preference [70,71].

#### Study design

The study will utilise an explorative, holistic, single-case study design with a flexible approach [68]. This design is considered most appropriate for addressing the study’s aim, as it allows for an in-depth investigation of an existing knowledge gap using multiple sources of evidence. Case-study methodology, as described by Yin [68], enables flexibility in framing and enquiring a specific phenomenon while maintaining a structured and rigorous approach. The case is defined as ‘Swedish young adults’ mental health and well-being during the transition to adulthood’. Furthermore, an inductive approach will be used to ensure that the findings remain as close as possible to the participants’ experiences and perspectives [68].

#### Theoretical study propositions

The theoretical study propositions, divided into ‘provisionary study propositions’ and ‘rival study propositions’ (see Table 1), were created by the first author (MKB) in discussion with AL and SG, reviewed by the co-authors (UJ, CH), and evaluated by an expert panel of young adults. They represent a limited selection of the possible explanations for the mental ill-health of Swedish young adults, stemming from the above-mentioned theories and perspectives included in the framework. The propositions are constructed to be distinct and different but not mutually exclusive. They will be used to guide the data analysis, triangulation and pattern-matching outlined in this protocol [68].

### Data collection procedures

The timeline of this case study is commenced and planned as following; recruitment of the various participant group started May 2024 and is planned until October 2025. The data collection with focus group interviews, individual interviews and documentation searches began August 2024 and will proceed until December 2025. Results are expected to be generated and reported in April 2026.

#### Sampling

According to the case study methodology outlined by Yin, ‘data saturation’ can be understood as and reached when the data collection has yielded sufficient evidence to test the study propositions [68]. More specifically for this study, data saturation is estimated to be reached with 4-6 focus group interviews per study participant group. Prior research on data saturation and its relationship with the number of focus group interviews needed has shown that 4-6 are sufficient for generating codes and subsequent themes covering the phenomena studied [74–76]. For this case study, we argue that the focus group interviews with the participant groups of stakeholders and parents will generate similar codes and subsequent themes and thus be summed to reach a total of 4-6 focus groups. Data saturation for the documentation of Swedish mental health and well-being promotion offered to young adults on municipal and regional levels is estimated to be reached when no new codes are generated from the material, labelled ‘coding saturation’ [77].

Participants for this case study have been or will be recruited using different purposive sampling techniques [78]. Firstly, 21 participants from relevant Swedish stakeholder organisations and groups were recruited through snowball and convenience sampling during May–June, 2024 [78]. The inclusion criteria were having worked a minimum of one year in the field of young adults’ mental health. Organisations contacted were local, regional, and national mental health and well-being services specifically targeting young adults, university counsellors, and non-governmental organisations. Potential participants received an email containing information about the study and participation.

Secondly, young adults have been and will be recruited trough maximum variation sampling [78] with paid advertisement campaigns on social media platforms and information about the study posted at libraries, gyms, pubs and clothing stores. Inclusion criteria are being between 18-29 years of age and speaking Swedish. The communication and strategy of such paid advertisement campaigns is done in partnership with a social media specialist at Lund University. The advertisements provide a brief introduction to the study, including a link to the clinical trials website hosted by Lund University’s medical faculty. The website contains detailed study information, participation details, and researcher contact information. After notifying their interest via e-mail or SMS, the young adults are asked to provide details regarding their living locality (countryside, small town, city), experiences with mental health/ill-health, and their current employment, education, or similar activity.

At the time of this case study protocol submission one round of recruiting young adults with paid social media campaigns had been performed during October–November 2024, with 57 young adults expressing interest in participating, and 20 having engaged in the focus group interviews. A second round of recruitment will commence in May 2025 focussing on recruiting participant groups underrepresented in the first round, primarily males between 18 and 29 and young adults identifying as LGBTQ+. Unlike to the first social media campaign, which featured images of young women, the second campaign will use images reflecting the targeted underrepresented groups. This sampling strategy has proved effective in previous mental health research [79].

Thirdly, parents of young adults aged 18 and 29 will be recruited using maximum variation sampling [78] and is planned to be initiated in May 2025. Inclusion criteria include being a parent to one or more young adults aged 18-29. Similar to the recruitment of young adults, parents of young adults will be recruited using paid advertisement campaigns on social media, in consultation with a social media specialist at Lund University. The advertisement will provide a brief introduction to the study and include a link to the clinical trials website hosted by Lund University’s medical faculty. The website will contain detailed study information, participation details, and researcher contact details. Interested parents will be referred to a survey hosted on a REDCap server [80], where they are asked to provide their name, age, email address, and a brief account of the motivations underlying their decision to participate.

#### Data collection

Focus group interviews will serve as the primary mode of data collection, given they are particularly suitable for exploring perspectives and interactions among participants [81]. An interview guide (see Appendix II) has been designed based on the research questions and tailored to the different participant groups attending the focus group interviews. The guide focuses on key knowledge areas, including the transition to adulthood, mental health literacy, and occupational balance. The questions are explorative and open-ended, for example: *“Can you share your experience of the transition to adulthood?”* and, *“What do you think is important for promoting mental health and well-being among young adults? Feel free to give examples”*.

During the planned focus group interviews [81], participants will respond and interact with certain interview questions using the online software ‘Mentimeter’ [82]. Responses will be displayed in real-time and discussed among the participants. Examples of questions using ‘Mentimeter’ [82] are, *“When considering everything you do in your daily life, how do you experience the balance between different activities?”* and, *”What characterises young adults’ transition to adulthood in your organisation?”*.

Phase 1: Focus group interviews (*k*=3, duration=120 minutes) and individual semi-structured interviews (*k*=2) were conducted with professionals belonging to stakeholder groups (*n*=22) in August and September 2024. Two individual semi-structured interviews were conducted with participants identified as information-rich cases, but were unable to attend the focus group interviews. A tailored interview guide ensured consistency across the focus group interviews and semi-structured interviews.

Phase 2: Focus group interviews (*k=*4, duration=120 minutes) with young adults (*n=*20) were conducted digitally during November and December 2024. A tailored version of the interview guide was used to ensure consistency across the focus group interviews. A second round of focus groups (*k*≈1-2, duration=120 minutes) is planned for May-June 2025 focussing on recruiting participants from previously underrepresented backgrounds ensuring we capture their perspectives on the transition to adulthood, mental health literacy and occupational balance.

Phase 3: Documentation [68] of existing preventive and mental health-promoting efforts and information provided to young adults across various municipalities and regions in Sweden will be mapped during October 2025. For this case-study, we have made a deliberate choice in researching prevention and promotion efforts rather than on the care and support available once someone has become ill. In Sweden, the responsibilities of providing mental health support and care are legally divided into four main pillars; 21 autonomous regional councils and 290 municipalities, with regional councils mainly providing emergency psychiatric treatments and rehabilitation for children, youth and adults, whereas the municipalities reside over psychosocial rehabilitation, prevention of mental ill-health and mental health and well-being promotion [83]. Employers and workplaces are responsible when mental ill-health arises at work, with universities or educational institutions providing mental health and well-being support to students [84,85].

Data will be collected through Google searches, using keywords such as “mental ill-health”, “mental health”, “mental well-being”, “mental health promotion” and “prevention of mental ill-health”, “anxiety”, “worry”, “stress” combined with “young adults” and various Swedish municipalities (e.g. “Stockholm” and “Kalix”). The selected municipalities will represent a diversity of population sizes, geographical locations, and sociodemographic characteristics. The search results are saved and complied into an Excel-document for further thematic analysis.

Phase 4: Two digital focus group interviews (duration=120 minutes) with parents (*n≈*12-16) to young adults are planned for May–June 2025 to complement the focus groups interviews with young adults and stakeholder groups. A tailored version of the interview guide will be used to ensure consistency across the focus group interviews.

#### Data analysis

The case study analysis will consist of four separate thematic analyses, [86] one for each type of data source. The data sources will be analysed chronologically, meaning that coding and generating themes and subthemes for one data source will be completed before analysis of the next begins. The thematic analyses [86] will be conducted by the research team, consisting of the authors and one young adult. In this case study, triangulation will be employed to enhance the credibility and depth of findings by integrating insights from multiple data sources [68].

The process will involve representatives from various stakeholder groups (young adults, parents, mental health professionals, non-governmental organisations, and researchers), comparing and consolidating themes derived across the different data sources. This approach ensures consistency and helps uncover diverse perspectives on the transition to adulthood and its impact on mental health, mental health literacy, and the occupational balance of Swedish young adults in Generation Z. Figure 1 provides an overview of the data collection process and planned analytical procedures.

**Figure 1.**
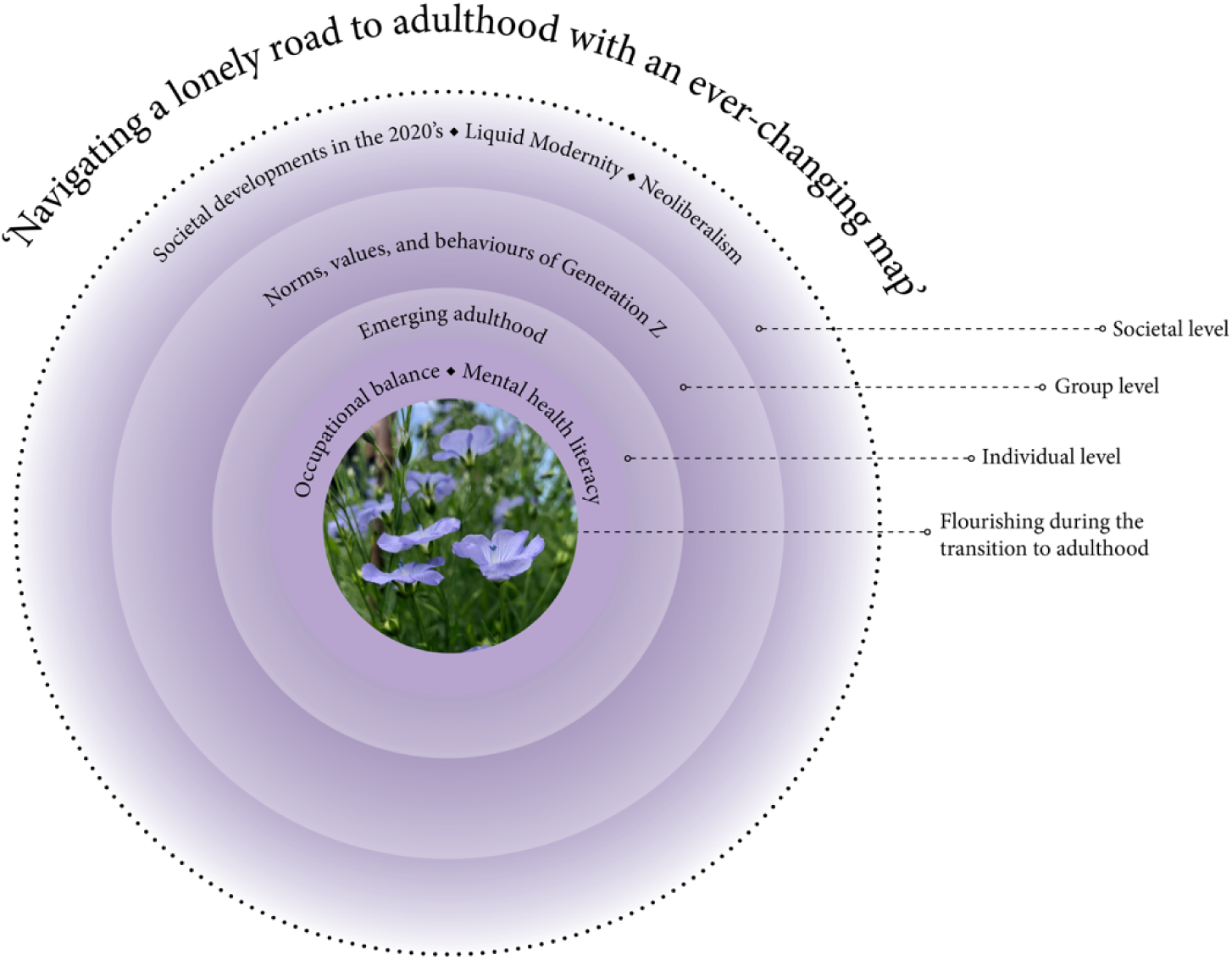
‘Navigating a lonely road to adulthood with an ever-changing map’: A framework for understanding today’s challenges to the mental well-being of Swedish young adults

**Figure 2.**
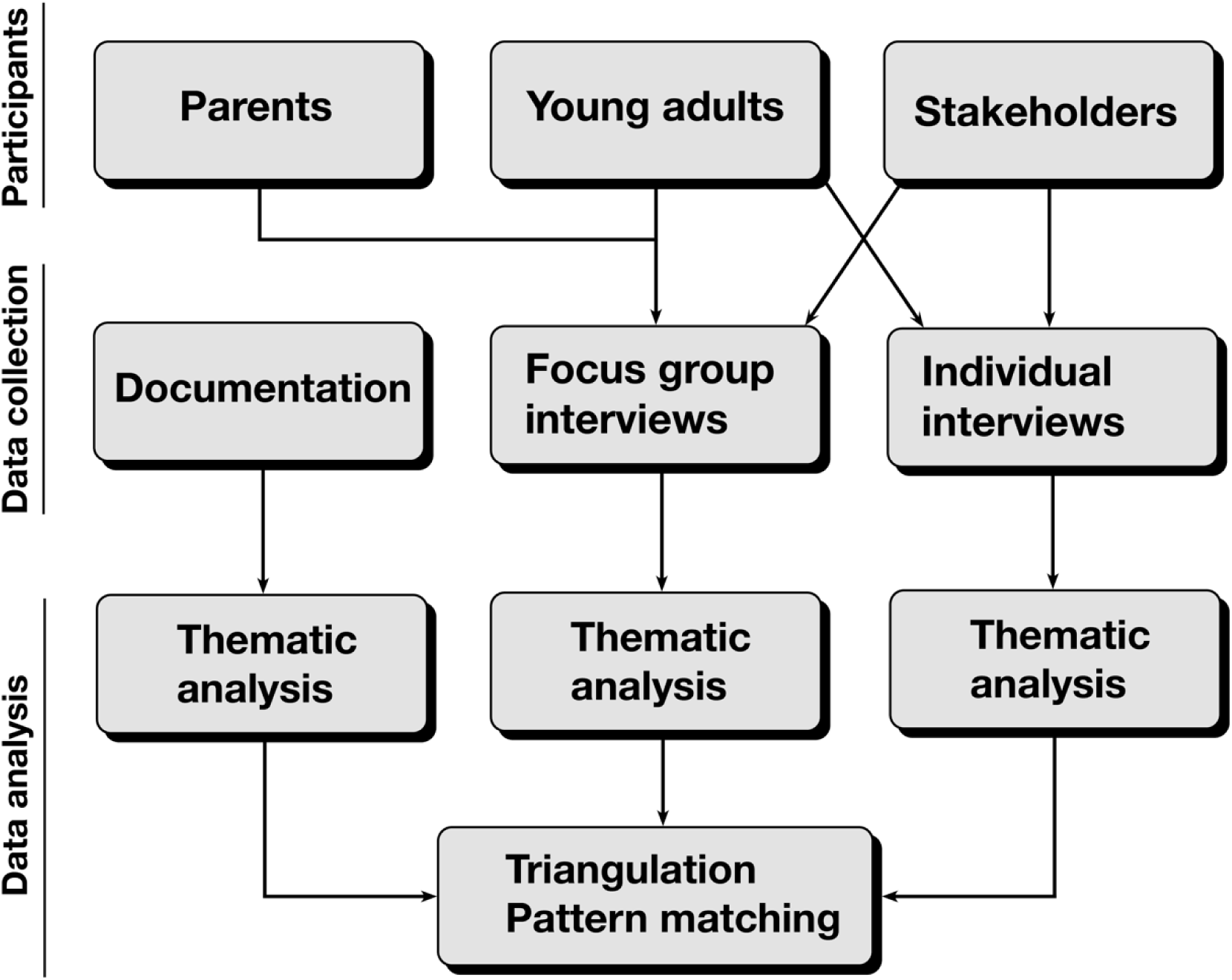
Diagram representing the planned data collection and analytic methods for a case-study investigating Swedish young adults’ (ages 18-29) mental health literacy, mental health and occupational balance during 2024-2025.

#### Thematic analysis

**Stage 1:** Each transcription will be read through for familiarisation. Initial codes will be created and applied to the segments of data relevant to the study aim. Each researcher conducting the coding will document personal reflections and thoughts after each coding session.

**Stage 2:** During a first data analysis conference, the researchers will meet to discuss the coding process, compare initial codes, and assess similarities and discrepancies. Based on these discussions, a set of revised codes will be developed for further abstraction and analysis.

**Stage 3:** The revised codes and data excerpts will be compiled into a structured document, together with suggestions, initial themes, and subthemes.

**Stage 4:** The research project’s expert group of young adults will review the revised codes, data excerpts, and drafts of themes and subthemes.

**Stage 5:** A second data analysis conference will be held with the research team and the young adult participant to refine themes and subthemes, ensuring their validity and alignment with data sets.

#### Triangulation and Pattern Matching

One workshop will be conducted with stakeholders (young adults, parents, mental health professionals, and representatives from non-governmental organisations, *n*≈10-12, duration=120 minutes) after each data source has undergone thematic analysis. The workshops will be interactive, using ‘Mentimeter’ [82] and breakout rooms in a digital video conferencing format. The two primary aims of this workshop are:

1. Triangulation of the data sources: The identified themes, subthemes, and interlinking chains of evidence [68] will be discussed and collectively evaluated through consensus-based decision making.
2. Pattern matching to case study propositions: The themes and subthemes will be analysed and mirrored against propositions A and B described in this research protocol, following Yin’s pattern matching approach [68].

The final situational analysis, which is the primary aim of this case study, will be shaped during a workshop with an expert panel of young adults (*n*≈9-10) aged 18-29. Collectively, they will discuss the initial themes and subthemes as well as the perspectives and opinions arising from the previous triangulation and pattern-matching workshop.

#### Processing and management of data

The data collected for this study is protected on a hard drive stored in a safe at the Faculty of Medicine, Lund University. Only two of the authors and analysts (MKB, AL) have the numerical code to access this safe. All interviews are recorded and thereafter saved as an audio file. Subsequent transcription is handled by a member of the research team gaining access to the audio files by transfer in person. The transcriptions are deidentified to ensure the study participants’ confidentiality. Sociodemographic data of the young adults and parents and their name initials are compiled in a list stored in the safe. According to Swedish law, the collected data and documents will be archived for 10 years after completion of the research project [87].

#### Use of artificial intelligence

According to the European Commission’s ‘Living guidelines on the responsible use of generative AI in research’ [88], researchers should demonstrate transparency in the usage of artificial intelligence. For this paper, generative artificial intelligence through OpenAI’s’ChatGPT’ [89] has been used during the writing process. Uses have been spell-checking, corrections on grammar and editing for language clarity. Further uses of ChatGPT have been to examine the logic of certain paragraphs and contents, discuss ideas and theories as well as searching for viable references.

#### Ethical considerations

The case study is approved by the Swedish Ethical Review Authority (ethical approval number 2024-00582-01), and adheres to the guidelines as outlined by the World Medical Association [90]. The participants received and will receive compensation in the form of a voucher connected to a supermarket and food store chain worth 100 SEK (approximately 9,12 €). The focus group interview as a qualitative method comes with certain ethical challenges, such as consent, knowing who they are but upholding confidentiality as well as the risk of harm [91]. Our strategies to minimise these risks are the following:

The research participant information letter has been approved by the Swedish Ethical Review Authority and was and will be handed out to participants prior any individual or focus group interviews. It entails a rich description of what a focus group interview or individual interview is, the time frame, eventual risks with participating and with which researchers it will be conducted. All participants have signed and will sign informed written consent prior or shortly after the interviews and are informed of their right to withdraw from the study at any time, as well as that their participation will be held confidential.

Keeping the confidentiality of study participants in the focus groups is addressed by setting ground rules in the beginning of each interview, as recommended by Sim and Waterfield [91], and described in each of the interview guides approved by the Swedish Ethical Review Authority. Firstly, all the participants are assured that their participation and what they share with the researchers will be deidentified; secondly, the participants are prompted to deidentify any persons if mentioned as examples; and thirdly promoting the message of,“What is said in the focus group stays in the focus group” is promoted to participants at the beginning of each session. The discussions are supervised by two researchers with clinical experience in mental health to ensure a safe and welcoming environment.

Given sharing one’s views or experiences of mental health and well-being in a focus group environment with strangers can be sensitive and potentially evoke feelings of anxiety, stress or fear (risk of harm) [91], available supports are outlined in the information letter. If focus group participants experience distressing feelings or thoughts following the interviews, they are encouraged to reach out to the last author (AL) who has many years of clinical experience working with young adults can advise on where to seek appropriate support. Should this initial support not be sufficient, participants may be referred to collaborating partners in mental health services.

Sufficient time is allowed both prior to and following each focus group interview to create a warm and welcoming safe space. Sharing emotionally charged experiences with strangers may be meaningful for participants when they regard the objectives of a study as relevant both on a personal level and for the broader community [92]. Accordingly, the overarching aims of the research project, along with its potential contributions to the mental well-being of young adults, are explicitly outlined in the information letter and repeated prior to each focus group interview.

## Discussion

### Methodological strengths and limitations

Conducting a case study is well-suited to study complex interplays concerning mental health and well-being needs and outcomes [93–95], due to its methodological emphasis on building a case with multiple chains of evidence and situating the research within the phenomena studied [96]. Another merit of case-study methodology lies in its balance between scientific rigour and its adaptability, allowing for adjusting the study design and data collection to better capturing the phenomena in its real-life context [68,96].

A common criticism of case studies is that they produce research with low levels of generalisability other than the specific context studied [68,96]. Responding to this piece of criticism, one of the primary aims and strengths of case study research is its emphasis on rich contextual descriptions, which is well suited to achieve theoretical generalisation [68,96].

This means to relate one’s study findings in developing, testing, and refining theories [68,96]. The unsuccessful prevention of mental ill-health and promotion of mental well-being among young adults warrants novel theories in order to understand and address the current mental health crisis [1,45]. Thus, we argue that a case study is well-opted in achieving this goal. We aim to bolster our case study’s theoretical generalisability through rich contextual descriptions of the context and case studied, triangulation of data sources, as well as actively co-analysing the material with a broad sample of participant groups, as recommended by Crowe et al. [96]. The theoretical generalisability will also be strengthened by the case study’s propositions building upon a wide selection of theories relevant for understanding the mental health of young adults, previously reviewed by an expert panel of young adults relating to their lived experiences [68,96].

## Conclusions

The urgent issue of mental ill-health among young adults calls for innovative efforts across society to promote their well-being. The framework ‘Navigating a lonely road to adulthood with an ever-changing map’ pinpoints the need of providing resources, knowledge and coping skills for young adults in this often-challenging life phase of emerging adulthood. At the same time, contemporary society is characterised by high levels of uncertainty and stressors further straining the mental health and well-being of young adults.

The situational analysis stemming from this case-study is expected to provide new insights into the factors driving the surge of mental ill-health among young adults. In particular, it aims to illuminate both the opportunities and challenges of emerging adulthood, as well as identifying the support needs of Swedish young adults’ in order to inform the development of future interventions targeting their mental well-being.

## Authors’ contributions

According to the Contributor Role Taxonomy (CRediT), the authors made the following contributions to the manuscript:

MKB: Conceptualization, Methodology, Project administration, Writing – original draft, Writing – review & editing, Visualization.

SG: Writing – review & editing, Funding acquisition, Supervision, Conceptualization, Methodology.

UJ: Writing – review & editing, Funding acquisition, Supervision. CH: Writing – review & editing, Funding acquisition.

AL: Writing – review & editing, Funding acquisition, Supervision, Conceptualization, Methodology.

## Data Availability

No data was produced for this study protocol.

## Acknowledgements

The authors want to express their sincere gratitude to the expert panel of young adults who provided their time, lived experiences and perspectives in co-creating the framework.

